# The symbiosis of healthcare professionals and patients: understanding the dynamics of long-term care relationships between healthcare professionals and patients in oncology

**DOI:** 10.1101/2024.08.13.24310782

**Authors:** Liam Il-Young Chung, Awranoos Ahadi, Grace Lee, Chan Mi Jung, Lena Chae, Ilene Hong, Grace Kang, Jessica Jang, Brandon Park, Young Kwang Chae

**Affiliations:** Northwestern University Feinberg School of Medicine, Chicago, IL; Northwestern University, Chicago, IL; University of Missouri-Kansas City School of Medicine, Kansas City, MO

## Abstract

**Background:** Chronic diseases are a significant source of physical, emotional, mental, and social distress to patients, caregivers, and healthcare professionals (HCPs). In particular, cancer patients face immense challenges as they navigate their uncertain futures. These difficulties necessitate a collaborative environment where patients, caregivers, and HCPs support each other to improve everyone’s quality of life. This study aims to investigate factors that influence a healthy long-term care relationship between patients and HCPs.

**Methods:** This qualitative study analyzes the experiences of the HCPs with patients and their caregivers at a large metropolitan academic medical center. Twenty-two de-identified reflective essays written by HCPs as part of the Pacemakers initiative at the medical center, which seeks to empower HCPs, patients, and caregivers in creative ways along the management journey were analyzed. The essays describe meaningful experiences, such as patient/caregiver award ceremonies celebrating resilience and hope. Essays (*N* = 22) were thematically analyzed by two independent coders to identify key codes and themes (italicized).

**Results:** The presence of *support networks* (*n* = 10) was crucial for patients and caregivers. *Conveying solidarity* (*n* = 21) helped patients experience companionship with their medical team, family, and friends. *Small acts of kindness* (*n* = 10) led to meaningful interactions that encouraged patients. *Compassion* (*n* = 19) and *patient-centeredness* (*n* = 17) fostered a receptive environment where patients and caregivers felt heard. Celebrating and honoring the *resilience* (*n* = 10) of the patients made them feel recognized, and this process of celebration was also found to bolster a *renewed sense of purpose for the HCPs* (*n* = 11).

**Conclusion:** The results highlight the importance of holistic support for patients and their caregivers. Creative encouragement empowers both patients and HCPs, building trust and strengthening the pursuit of their health. This mutual care fosters an environment conducive to healing and recovery. This study offers insights into effectively supporting patients and HCPs in long-term care relationships.

## Introduction

Cancer has profound psychological, emotional, and social effects on patients, caregivers, and healthcare professionals (HCPs). Cancer patients often suffer emotionally, experiencing depression and anxiety, which can worsen their condition, adherence to prescribed treatments, and quality of life [1,2]. As most cancer treatments occur in outpatient settings, patients are often found in a position where they must take an active role in managing their health outside of the hospital, which involves significant lifestyle changes and symptom management [1,3,4,5]. Numerous studies highlight the financial and psychological toll of cancer treatment on patients and their caregivers [6,7]. Employed cancer patients experience additional distress, as their ability to work may be affected, leading to additional financial and emotional burdens [8].

Informal caregivers, often friends or family, play pivotal roles in patients’ lives by managing practical responsibilities and prioritizing patients’ well-being [9,10,11]. Yet, caregivers also experience emotional distress and declining well-being. Depression, anxiety, and somatization are commonly experienced challenges by caregivers [2,12,13], emphasizing the need for robust support systems tailored to caregivers’ needs. Despite the challenges, caregivers are vital to patient outcomes, as strong social support can improve patient health and survival [14,15,16].

HCPs (physicians, nurses, and allied health professionals) face significant challenges as well, leading to burnout characterized by emotional exhaustion, depersonalization, and a diminished sense of accomplishment, with prevalence rates exceeding 50% in the United States [17,18]. HCP burnout jeopardizes patient safety, care quality, and patient health outcomes, emphasizing the necessity of addressing burnout in healthcare [19].

Given these challenges, fostering supportive relationships between HCPs, patients, and caregivers emerges as a pivotal strategy in alleviating the burdens associated with cancer treatment. One strategy to combat burnout and overall distress involves nurturing long-term care relationships between HCPs, patients, and caregivers. Patients value physician empathy, trust, and competence in these relationships [20]. Such relationships can not only alleviate the burdens on everyone involved but also positively influence the psychosocial dynamics of patient care. This study aims to explore factors that contribute to the cultivation of supportive long-term care relationships and alleviate the burdens of cancer treatment, ultimately influencing health outcomes.

## Methods

### Study Design and Sample

This study qualitatively examined de-identified reflective essays of twenty-two HCPs who collaboratively interacted with patients and caregivers in the clinical setting through the Pacemakers initiative, implemented at a large urban academic medical center. The initiative aimed to foster creative encouragement and companionship among HCPs, patients, and caregivers. As part of this initiative, HCPs held ceremonies with personal awards for patients and caregivers during their treatment course. HCPs were given the option to submit de-identified reflection essays as part of a mindfulness campaign and team-building exercise. Thematic analysis was performed on these essays to identify recurring themes in HCPs’ experiences.

### Analysis

Two objective coders independently analyzed the data, following Braun and Clarke’s six-step thematic analysis guideline [21]. The coders first actively and thoroughly read the essays (*familiarizing yourself with data*). Afterward, potential data items of interest, questions, connections between data items, and other preliminary ideas were identified to generate codes, which are defined as the “most basic segment, or element, of the raw data or information that can be assessed in a meaningful way regarding the phenomenon” [22] (*generating initial codes*). From these codes, the coders searched for potential themes of broader significance (*searching for themes*). Next, the codes and themes were re-examined and re-sorted at the discretion of the coders (*reviewing themes*). Following this recursive step, the coders created definitions and narrative descriptions for the themes while identifying emergent sub-themes as necessary (*defining and naming themes*). Lastly, the data were organized and reported based on the analysis conducted through these steps (*producing the report/manuscript*).

### Results

A total of twenty-two de-identified essays from HCPs were analyzed. The following themes were identified as having the most impact on those involved in cancer treatment. Conveying *solidarity* (*n* = 21) was essential in fostering a sense of companionship and fellowship within their medical team, family, and friends. This sense of solidarity helped patients feel supported and connected. Additionally, small acts of kindness, which are often perceived as “*little means much*” (n = 10), conveyed genuine interest in patients and led to encouraging and meaningful interactions. *Compassion* (*n* = 19) and *patient-centeredness* (*n* = 17) created a receptive environment where patients and caregivers felt heard. Celebrating patients’ *resilience* (*n* = 10) made patients feel recognized and gave a *renewed sense of purpose for the HCPs* (*n* = 11). *Support networks* (*n* = 10) were identified as crucial for patients and caregivers, providing essential emotional and practical assistance.

HCPs *benefitted* from participating in creative patient encouragement ceremonies, which were found to be *motivating* or *inspiring* (*n* = 15), helping to prevent burnout. HCPs *felt acknowledged* for their support (*n* = 5) and appreciated the *opportunity to acknowledge their patients and caregivers* (*n* = 13). Additionally, HCPs were able to *connect emotionally* (*n* = 14) with patients and caregivers through shared laughter and tears, fostering a sense of *togetherness* (*n* = 13) and a more *personal connection* (*n* = 15) with patients and caregivers through these ceremonies.

## Discussion

### Patients’ Perspectives

#### Social Support Networks

Patients (10 out of 22) report that social support networks for patients, including family, friends, and colleagues, are crucial for providing practical, emotional, and financial assistance, thereby fortifying resilience throughout their journey. This support extends beyond emotional and physical aspects, aiding patients in adhering to treatment plans and making necessary lifestyle changes that influence positive outcomes.

In oncology where patients’ lives are burdened by prognosis and treatments, a healthy relationship between the patient and HCP underscores the significance of emotional support that can come from the HCPs and the patient’s family and friends, thus influencing the patient’s choice and strength to adhere to rigid treatment protocols. One essay highlights an encounter between an HCP and a patient who, despite receiving distressing news about her disease, remained hopeful about future treatment plans due to a strong relationship with her HCP. This encounter emphasizes the importance of the patient-provider relationship and the firm trust patients place in their HCPs to help them make decisions that align with their values, including choosing between continuing a difficult treatment plan or prioritizing their quality of life.

However, it is not just the patient-provider relationship that influences patients’ decisions to continue or discontinue treatment but also the patients’ social circle, specifically friends and family. These individuals serve as pillars of strength, helping patients navigate the complexities of their healthcare journeys, especially amidst significant emotional and physical pain. Another HCP in our study highlighted the crucial role played by a patient’s guardian during hospital visits and treatment appointments. This accentuates the fact that patients derive immense emotional and practical support from their close social networks, which substantially helps them address the challenges of undergoing cancer treatment.

#### Little Means Much

The concept of “little means much” underscores the significant impact that daily, small acts of kindness and brief positive interactions can have in patient care. Patients (10 out of 22) reported that these seemingly minor gestures, such as positive updates and daily words of encouragement, hold immense value for both patients and caregivers. For patients, these interactions can serve as a constant source of hope and positivity, boosting their mood and motivating them to adhere to treatment plans. For HCPs, these small acts provide emotional support and help maintain a positive outlook, enabling them to deliver optimal care. Thus, fostering a supportive and encouraging environment through these small acts can contribute to improved long-term outcomes for patients.

#### Recognizing Resilience, Not Just “Success”

In patient care, acknowledging the resilience of individuals facing cancer rather than solely focusing on treatment outcomes allows patients to feel empowered despite their prognosis. Several patients (10 out of 22) reported that appreciation for their efforts encourages them to persevere through treatment. Even without a favorable prognosis, patients often express gratitude from words of encouragement from HCPs and feel more confident in making informed treatment decisions.

Our study highlights three compelling examples of patients uplifted by such acknowledgments. Despite their condition, one patient expressed gratitude for each day, celebrating the present moment. Another patient, who received the “Milestone Award,” was inspired to persevere through difficulties after hearing encouraging words and appreciation from HCPs. The third patient was deeply moved by the “Sunshine Award,” which was adorned with encouraging messages.

Recognizing and valuing patient resilience is vital in compassionate healthcare. Acknowledging patients’ efforts empowers them, providing motivation and comfort amid uncertainty and physical and emotional pain. Prioritizing resilience alongside treatment outcomes fosters a holistic and compassionate approach to cancer care. This approach not only supports patients in their journey but also enhances the overall patient experience, promoting a more humane and supportive healthcare environment.

#### Compassion of Healthcare Providers

The manner in which news is delivered to the patient by HCPs significantly influences the patient’s emotional state and sets the tone for the rest of the treatment plan and the patient’s adherence to it. A majority of the patients (19 out of 22) expressed the vital role of compassion from HCPs for the patient’s well-being, both short- and long-term. Demonstrating genuine compassion to address patients’ distress allows patients to regain a sense of independence and dignity.

However, HCPs are seldom alone in such situations. While a single HCP can make a meaningful impact, a team approach creates a more sustainable positive environment. Therefore, the focus should shift to healthcare teams rather than a single HCP. Compassion is crucial when delivering distressing and difficult news, and all members of the healthcare team must exhibit it equally to sustain emotional support. Examples from our study show how HCPs engaging with their patients’ interests and stories can provide a sense of comfort, making the overall treatment experience less challenging.

Thus, HCPs’ compassion is a cornerstone of exceptional patient care, significantly shaping a patient’s emotional well-being and the cancer treatment journey. It empowers patients, offers emotional support, and builds trust. Compassion extends beyond medical facts, fostering comfort and trust. Its profound impact underscores its essential role in holistic healthcare, positively contributing to patient outcomes and well-being.

### Healthcare Providers’ Perspectives

#### Emotional Connection with Patients/Caregivers

While empathy is most often contextualized from the patient’s perspective, our study illuminates the positive impact of such an emotional connection on the empathizers—the HCPs—themselves, and their cross-influence on patient care and treatment outcomes.

In our sample of 22 HCPs, 14 elaborated on feeling a strong emotional connection with patients and caregivers. One HCP described, “It also brought a trembling of emotion to my heart,” upon witnessing their patient’s emotional response to an award ceremony. They added, “Even though I couldn’t [completely] hear [the patient’s] response, I was still able to laugh with them happily.” Such accounts demonstrate how empathetic displays of encouragement also affect caregivers, as shown by another HCP’s comments: “It creates a mysterious resonance that comforts both the receiver and the giver.”

With empathy evident as a key pillar for quality care, concerns arise about the emotional toll it may exert on HCPs. A more traditional approach to patient interaction emphasizes rationality over compassion. It is essential to find a balance between empathy and emotional detachment, which is intriguing to explore in the context of the creative encouragement process.

#### More About HCP Burnout

Physician burnout has become a prevalent issue in healthcare, driven by demanding schedules, perpetual workloads, and the emotional toll of delivering distressing news to patients. This burnout can negatively impact the quality of patient care and safety. Hence, addressing the emotional and physical challenges faced by HCPs is crucial for improving healthcare quality.

In our analysis, one HCP described the experience as “emotionally taxing” when witnessing patients’ deteriorating conditions. However, the establishment of enduring, meaningful relationships with patients was noted to alleviate some of this emotional burden. These connections not only bring comfort to patients but also reinforce the potential of treatments for better outcomes. Positive patient-provider relationships play a crucial role in mitigating physician burnout and fostering a healthier environment overall.

#### Personal Connection with Patients/Caregivers

Establishing personal connections between HCPs and their patients, including caregivers, proves to be vital and transformative, as evidenced by our analysis of twenty-two HCPs. These connections result in deeper relationships with patients and their caregivers, fostering mutual trust and understanding.

Six HCPs provided detailed accounts of forming personal bonds through meaningful conversations about various aspects of the patient’s life, including health, lifestyle, culture, and beliefs. Two HCPs recounted instances where patients openly expressed their fears about their illnesses and treatment, which led to increased empathy and a commitment to enhanced care. Additionally, four HCPs recalled gaining insights into patients’ lives beyond their illnesses, which enriched their understanding of patients’ diverse backgrounds and inspired cultural competence in patient care for better rapport with patients.

Another HCP shared a heartwarming encounter where a joyful spiritual conversation with a patient culminated in a heartfelt hug, demonstrating the profound trust and connection between HCPs and patients.

While maintaining objectivity is essential for optimal care, these meaningful interactions build mutual trust and enable HCPs to align treatments with patient values. They bring joy and motivation to HCPs and deepen their investment in patient well-being. Nurturing these personal connections is thus fundamental to achieving patient-centered healthcare.

#### Feeling Acknowledged / Being Able to Acknowledge Patients

Our qualitative analysis reveals the extensive impact of creative patient encouragement ceremonies on the overall wellness of HCPs. These ceremonies provided a unique platform for mutual acknowledgment between HCPs and their patients, reinforcing their dedication and commitment to the medical field.

Among the twenty-two HCPs sampled, one recounted a heartwarming moment when a patient received a “Milestone Award” after a three-year chemotherapy course. Witnessing the entire team celebrate the patient’s strength and resilience left an indelible mark on this HCP, who developed a deep admiration for the fortitude displayed by cancer patients.

Another HCP shared a touching instance where a patient received the “Sunshine Award” along with heartfelt messages from their entire care team. In response, the patient expressed gratitude for their care team, solidifying the bond between HCPs and patients through mutual gratitude and appreciation.

Furthermore, two HCPs emphasized the significance of feeling acknowledged by patients during hospital visits, highlighting the mutual trust that sustains HCPs in their duties and commitments. This recognition and trust encouraged HCPs to consistently deliver high-quality care.

While patient encouragement ceremonies primarily aim to uplift patients during physically and emotionally challenging journeys, our findings underscore that these ceremonies equally enhance the wellness of HCPs. By reinforcing their dedication to their profession, these ceremonies remind HCPs of their initial motivations for entering the field. Strengthening the bond between patients and HCPs is a pivotal goal of these creative patient encouragement ceremonies, achieved through mutual trust and acknowledgment, ultimately benefiting all stakeholders involved in cancer treatment and contributing to improved health outcomes.

#### Renewed Sense of Purpose

Creative patient encouragement ceremonies not only bolster patient resilience during cancer treatment but also positively impact HCPs. These ceremonies foster mutual admiration between HCPs and patients, nurturing long-term relationships and providing HCPs with a renewed sense of purpose and motivation.

A substantial majority of HCPs (14 out of 22) reported that patient interactions transformed their professional outlook, reminding them of the importance of their roles in patients’ lives. This revitalized purpose extended beyond healthcare, driving their dedication to ongoing education and research.

Patients’ unwavering positivity serves as a beacon of hope. Notably, seven HCPs highlighted the role of patient positivity during treatment. Acts such as wearing affirmational shirts or displaying Bible verses demonstrated patients’ resilience, profoundly impacting HCPs by rejuvenating their energy and hope, especially when delivering difficult news.

Moreover, seven HCPs reflected on “finding positivity in the dark” when delivering unfavorable news to patients. These challenging moments deepened their appreciation for their roles, highlighting the impact of their expertise and research on patient outcomes. Despite these challenges, HCPs found solace and gratitude, reinforcing their sense of purpose.

While patient-provider relationships are pivotal in bolstering patient strength during cancer treatments, our findings underscore the critical role of creative patient encouragement and robust patient-provider relationships in enhancing HCP wellness. Patient positivity and resilience significantly influence HCPs’ professional perspectives, emphasizing the importance of cultivating positive patient-provider interactions and celebrating patient resilience, which contribute to the well-being and motivation of HCPs.

## Conclusion

Cancer treatment profoundly affects the physical, psychological, and emotional health of patients, caregivers, and HCPs. Patients face lifestyle changes and uncertainties, while caregivers also experience significant challenges. HCPs can suffer from burnout, impacting the quality and safety of care provided.

This qualitative study highlights the challenging nature of cancer treatment and disease progression, underscoring its potential to lead to burnout among HCPs, thereby affecting patient care and the well-being of all involved. However, fostering long-term personal and spiritual care relationships between patients, caregivers, and HCPs can ease this burden. These relationships provide channels to navigate the challenges of cancer treatment by fostering deep emotional connections. Recognizing the importance of these connections not only improves overall well-being but also enhances outcomes and the quality of care for those affected

## Data Availability

All data produced in the present study are available upon reasonable request to the authors.

## Statements and Declarations

### Author contribution

The study conception and design were contributions from Liam Il-Young Chung, Grace Lee, Lena Chae, and Young Chae. Liam Il-Young Chung and Grace Lee conducted the study, synthesized information, and organized the content. The initial draft of the manuscript was prepared by Liam Il-Young Chung, Awranoos Ahadi, Grace Lee, Chan Mi Jung, Ilene Hong, Grace Kang, and Jessica Jang. Editing and revision were carried out by Liam Il-Young Chung, Awranoos Ahadi, and Brandon Park. Oversight of all steps was provided by Young Kwang Chae. All authors provided feedback on earlier manuscript versions and participated in the approval of the final manuscript.

